# Genetic Factors Associated with Suicidal Behaviors and Alcohol Use Disorders in an American Indian Population

**DOI:** 10.1101/2023.05.12.23289926

**Authors:** Qian Peng, David A. Gilder, Rebecca Bernert, Katherine J. Karriker-Jaffe, Cindy L. Ehlers

**Author notes:** Corresponding Author: Qian Peng, The Scripps Research Institute, 10550 North Torrey Pines Road, La Jolla, CA 92037 USA.

## Abstract

American Indians (AI) demonstrate the highest rates of both suicidal behaviors (SB) and alcohol use disorders (AUD) among all ethnic groups in the US. Rates of suicide and AUD vary substantially between tribal groups and across different geographical regions, underscoring a need to delineate more specific risk and resilience factors. Using data from over 740 AI living within eight contiguous reservations, we assessed genetic risk factors for SB by investigating: (1) possible genetic overlap with AUD, and (2) impacts of rare and low frequency genomic variants. Suicidal behaviors included lifetime history of suicidal thoughts and acts, including verified suicide deaths, scored using a ranking variable for the SB phenotype (range 0-4). We identified five loci significantly associated with SB and AUD, two of which are intergenic and three intronic on genes *AACSP1*, *ANK1*, and *FBXO11*. Nonsynonymous rare mutations in four genes including *SERPINF1* (PEDF), *ZNF30*, *CD34*, and *SLC5A9*, and non-intronic rare mutations in genes *OPRD1*, *HSD17B3* and one lincRNA were significantly associated with SB. One identified pathway related to hypoxia-inducible factor (HIF) regulation, whose 83 nonsynonymous rare variants on 10 genes were significantly linked to SB as well. Four additional genes, and two pathways related to vasopressin-regulated water metabolism and cellular hexose transport, also were strongly associated with SB. This study represents the first investigation of genetic factors for SB in an American Indian population that has high risk for suicide. Our study suggests that bivariate association analysis between comorbid disorders can increase statistical power; and rare variant analysis in a high-risk population enabled by whole-genome sequencing has the potential to identify novel genetic factors. Although such findings may be population specific, rare functional mutations relating to PEDF and HIF regulation align with past reports and suggest a biological mechanism for suicide risk and a potential therapeutic target for intervention.

## 1. INTRODUCTION

Suicide is a preventable public health problem that ranks as the second leading cause of death among young adults residing in the US (1). Suicide rates remain even higher among American Indians/Alaska Natives (AI/AN), with rates more than 50% greater when compared to the general U.S. population across all age groups (2–5). While suicide rates are unquestionably higher in the AI/AN population as a whole as compared to other US populations, rates also vary based on geographic region, tribal affiliation and whether one is living on a reservation (6–12).

American Indian/Alaska Natives also suffer from a disproportionate burden of the effects of alcohol, tobacco, and drug dependence (13, 14). Large-scale U.S. epidemiological studies indicate that, compared with other U.S. ethnic groups, AI /AN demonstrate the highest rates of alcohol and other drug dependence (15, 16). Additionally, lifetime rates of alcohol use disorders (AUD) differ depending on the tribes evaluated. Among individual tribal groups studied, reported rates of AUD have ranged from 20% to 70% (17–20)—significantly higher than epidemiological rates of AUD (14%) in the U.S. general population (21).

Substantial comorbidity has been demonstrated between SB and AUD. Chronic alcohol use is a clear risk factor for suicide (22), and having a severe use disorder and experiencing substance associated depression is particularly linked to increased risk for suicide (23). Being acutely intoxicated provides additional risk for lethal suicide over and above the risk of chronic use (24, 25). This is particularly true among American Indian/Alaska Natives (25, 26). However, documenting comorbidity between disorders does not necessarily imply a causal connection between them or a common etiological pathway. Both AUD and suicide have been shown to have a significant genetic component to their etiology (27, 28). Behavioral genetics studies have the advantage of being one of the strongest methods for determining whether the comorbidity among psychopathological conditions may be due to shared etiologies and/or pathologies associated with the disorders. For instance, a new study recently estimated the genetic correlation between suicide attempt and alcohol dependence at 44% in populations of European ancestry (29).

In recent years, numerous large genome-wide association studies (GWAS) have been reported for suicidal behaviors (28, 30–34). These studies were carried out in populations dominated by individuals of European descent. Genetic epidemiology studies have estimated the heritability of SB ranging from 17% to 55% (35, 36). Single nucleotide polymorphism (SNP)-heritability of suicide attempt using common genomic variants across the genome has been estimated to range from 3.5% in the UKBiobank (37) to 6.8% in the International Suicide Genetics Consortium (ISGC) (32). The largest GWAS meta-analysis to date (with ∼959K individuals) reported 12 loci for suicide attempt. Risk loci were mostly intergenic and implicated genes *DRD2, SLC6A9, FURIN, NLGN1, SOX5, PDE4B*, and *CACNG2* (30). Intriguingly, one of the genes, *ROBO2*, reported by the ISGC GWAS to be associated with SB (31) is on a rare run-of-homozygosity (ROH) segment that was previously reported to be linked to severe AUD in an American Indian population (38).

While SNP-heritability and GWAS studies primarily estimate the impacts of common genomic variants, recent studies have shown that rare variants, especially those in low linkage disequilibrium (LD) with neighboring variants, are enriched for heritability for complex traits and diseases. Rare variants often represent recent and potentially deleterious mutations that can have biological consequences. Studies have indeed shown that most rare missense alleles in humans are deleterious (39). In fact, a recent Utah Suicide Genetic Risk Study (USGRS) interrogating rare protein-coding variants included on the Illumina PsychArray chip has identified five rare variants for suicide death (40). Genotyping chips, however, are usually not designed to capture rare variants in a population. Thus, to facilitate comprehensive studies of the impact of rare variants on a disease, exome sequencing or whole-genome sequencing is usually necessary.

The present report is part of a larger study exploring risk factors for substance dependence and suicide behaviors among American Indians (19, 41–43). This American Indian (AI) population has been sequenced and deep phenotyped. The lifetime prevalence of AUD and suicide in this AI population is high, and evidence for heritability, linkage to specific chromosome locations, and genome-wide findings for AUD have been demonstrated (38, 42, 44-47). The present study aimed to evaluate genetic factors associated with SB in this AI population. We hypothesized that certain genetic variants may underlie both SB and AUD, and further, that joint association analysis of related traits may lead to increased power to detect variants that contribute to both phenotypes. To this end, we conducted a bivariate genome association analysis between SB and AUD. We further hypothesized that rare variants in a high-risk population maybe a significant component of the complex genetic architecture underlying the disorder. We thus conducted gene-based and pathway-based rare and low-frequency variant analyses for SB.

## 2. MATERIAL AND METHOD

### 2.1 Participants

American Indian participants were recruited from eight geographically contiguous reservations with a total population of about 3,000 individuals. To be included in the study, participants had to be between the ages of 18 and 70 years, and mobile enough to be transported from their home to The Scripps Research Institute (TSRI). More details are given in Supplemental Materials. The protocol for the study was approved by the Institutional Review Board (IRB) of TSRI, and the board of the Indian Health Council, a tribal review group overseeing health issues for the reservations where the recruitment was undertaken. Written informed consent was obtained from each participant after the study was fully explained.

### 2.2 Phenotypes and genotypes

Potential participants first met individually with research staff, and during a screening period, participants completed a questionnaire that was used to gather information on demographics, personal medical history, and drinking history (48). Each participant also completed an interview based on the Semi-Structured Assessment for the Genetics of Alcoholism (SSAGA) (49), which was used to collect lifetime history of two types of self-directed violence: suicidal thoughts including ideation (Have you ever thought about killing yourself?) and/or plans (Did you have a plan? Did you actually consider a way to take your life? What were you going to do?), as well as suicidal acts, including suicide attempt history (Have you ever tried to kill yourself? How did you try to kill yourself?) reported by the participants and suicide deaths obtained from community sources (e.g., verified by public records, family/tribal informants) over an 8 year period. From the lifetime history of suicidal thoughts and acts, we defined the suicidal behavior phenotype as a ranking variable: 0-none, 1-ideation, 2-plans, 3-attempts, 4-death.

Diagnoses of lifetime DSM-5 AUD (mild, moderate, or severe) were also generated using the SSAGA. In addition, the interview retrospectively asks about the occurrence of alcohol-related life events, and the age at which the problem first occurred, from which a quantitative phenotype, the severity level of AUD, was derived. Briefly, it is indexed by 36 alcohol-related life events (Table S1) in the clinical course of the disorder (19, 50), with life events given a severity weight of 1 for events 1-12; 2 for 13-24; and 3 for 25-36. AUD severity was then calculated as the sum of the severity weights of the 36 life events (46). The relation between the SB phenotype and the AUD severity in this AI population is illustrated in Supplemental Figure S1. The Pearson’s correlation between SB and AUD severity is 0.3, with a 95% confidence interval of 0.23-0.36.

Participants had low-coverage whole genome sequencing on blood-derived DNA using a previously described pipeline (51). Further details can be found in the Supplementary Method.

### 2.3 Bivariate association analysis for suicidal behaviors and alcohol use disorders (SB-AUD)

We conducted genome-wide bivariate association analysis to identify genetic variants that might be associated with both SB and AUD severity (denoted as SB-AUD). By leveraging cross-trait covariance, multivariate tests for association may provide increased power over univariate tests. This occurs when the residual correlation between traits is opposite in direction to the genetic correlation induced by the genetic loci (52). To control for population admixture and familial relatedness, we used the multivariate linear mixed model implemented as *genome-wide efficient mixed-model association* (GEMMA) for the study (53). The bivariate association for each variant was conditioned on a genetic relationship matrix of the cohort derived from the genotypes, thus capturing a wide range of sample structures. Gender, age, and age-squared were further included as covariates. Variants with minor allele frequency (MAF) lower than 1% were excluded from this analysis.

### 2.4 Gene-based rare and low-frequency variant analysis for suicidal behaviors

We included both rare (MAF < 1%) and low-frequency (1% <= MAF < 5%) variants in a second set of analyses. We use the term “rare variants” broadly in this text to refer to variants with MAF < 5%. Rare variants are usually tested by aggregating them into groups. We analyzed the rare and low-frequency variants across the genome using SKAT-O that optimally combines a burden test and a non-burden sequence kernel association test (SKAT) (54) within a linear mixed model as implemented in EPACTS (55). The variants were grouped by genes or by pathways and tested against the SB phenotype. Gender, age, and age-squared were included as covariates in all sets of analyses.

For each gene, we formed two types of groups. One group considered all variants on exons, 5’ and 3’ untranslated regions (UTRs), upstream and downstream of the gene (denoted as *ExonReg*). The other group included only the nonsynonymous variants and the splicing-site variants of the gene (denoted as *Nonsyn*). Intergenic and intronic variants were excluded in the present study. For each group type, a gene was excluded if fewer than three markers were found, or if less than 0.5% of the samples had any such markers on the gene. This resulted in 28,718 genes in the *ExonReg* group and 12,588 genes in the *Nonsyn* group. Association analysis was performed between each gene-based set and SB. False discovery rates (FDR) controlled by the Benjamini–Hochberg procedure (Benjamini and Hochberg, 1995) were used to set significant *p* values from the test statistics of the association tests. We combined two gene variant groups together for the multiple testing correction and report the FDR-adjusted *p* values (Yekutieli and Benjamini, 1999). Note that the correction is conservative since the *Nonsyn* variants on a gene are a subset of the *ExonReg* variants, and thus they are correlated.

### 2.5 Pathway-based rare and low-frequency variant analysis for suicidal behaviors

We utilized a collection of canonical pathways from the Molecular Signatures Database (MSigDB) version 7.5.1 (56, 57). There are total of 2,981 pathway gene sets in this release. Since a pathway may contain a large number of genes, to limit the number of variants included in each set for test, we only considered rare and low-frequency nonsynonymous and splicing-site variants (*Nonsyn*) on the genes within each pathway. The same filters as in the gene-based tests were applied. We performed SKAT-O tests on the *Nonsyn* variant group for each pathway to be associated with SB. FDR-adjusted *p* values are reported.

### 2.6 Functional Analysis

For the top variants identified in the bivariate association analysis and for the top genes identified in the rare variant analysis, we obtained the combined annotation dependent depletion (CADD v1.6) scores (58) for the variants included in the test to assess the deleteriousness of these variants. CADD integrates multiple functional annotations and genome-wide variant effect prediction models to produce a scaled C-score. We report the number of variants tested in each gene that have C-scores in the range of 15-20 (1-3% most deleterious mutations), 20-30 (0.1-1%), and over 30 (<0.1%). We chose 15 as the lowest C-score to report as it happened to be the median value for all possible canonical splice site changes and nonsynonymous variants in CADD v1.0 (59).

The variants with *p*-value < 10^-6^ from the bivariate analysis were annotated with genes, and the associated set of genes was then subjected to functional enrichment analysis using GENE2FUNC in FUMA version 1.5.1 (60). An integrated network analysis was completed using GeneMANIA (61). The same set of functional analysis was applied to the top genes from the rare variants analysis as well.

More details can be found in Supplemental Method.

## 3. RESULTS

A total of 743 participants had sequencing data and a non-missing phenotype for SB (*none [0], suicidal ideation [1], suicide planning [2], suicide attempt [3], suicide death [4]*), and 742 participants had data on AUD severity (range 0-69, mean=22.6, sd=18.8).

### 3.1 Bivariate genome-wide significant variants associated with SB-AUD in AI

Five variants were identified as being significantly associated with AUD severity and SB (*p*<5E-8), as illustrated in Figure 1. Only one of the variants, rs184204326 on gene *FBXO11*, has MAF over 5% (Table 1). Although the association of this variant is mostly driven by SB (*p*=1.48E-07), the bivariate association is statistically more significant (*p*=3.63E-08) than the univariate associations, which also holds for three other top variants, including an intergenic variant between genes *ZIC2* and *PCCA*, variant rs76300969 on gene *AACSL*, and variant rs530542541 on *ANK1*. Among the top variants shown in Table 1, rs184204326 is the only SNP for which brain expression quantitative trait loci (eQTL) were found in the Braineac database (62). That variant is associated with the differential gene expression of several genes in brain tissues including: putamen, substantia nigra, thalamus, occipital cortex, and temporal cortex (Table S2). The most significant eQTL is for *EPCAM* (encoding epithelial cell adhesion molecule) expression in the putamen tissue (*p*=0.0019). The variant is also associated with gene expression in several brain tissues for *FBXO11*, *MCFD2* (encoding a soluble luminal protein), and *KCNK12* (encoding a potassium channel protein).

**Figure 1.**
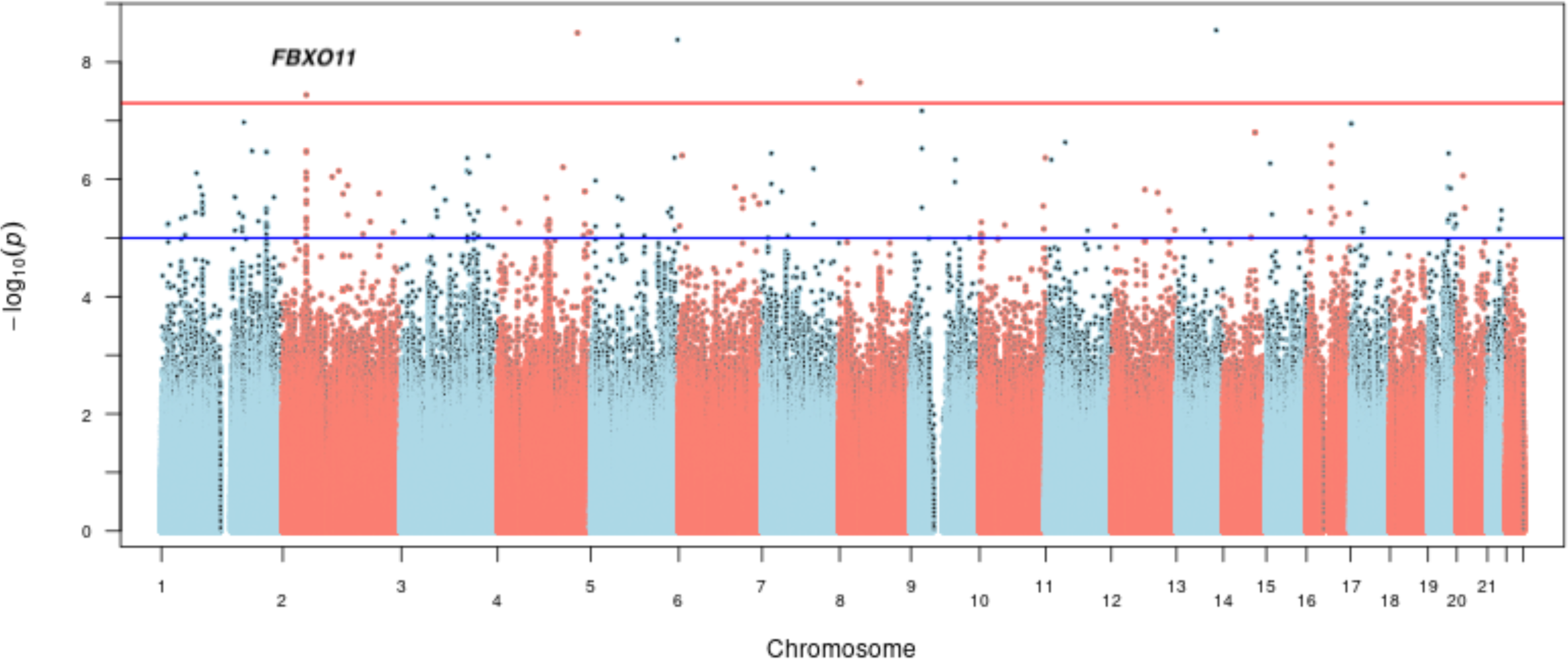
Manhattan plot of bivariate genome-wide association analysis for AUD severity and suicidal behaviors in AI.

**Table 1.**
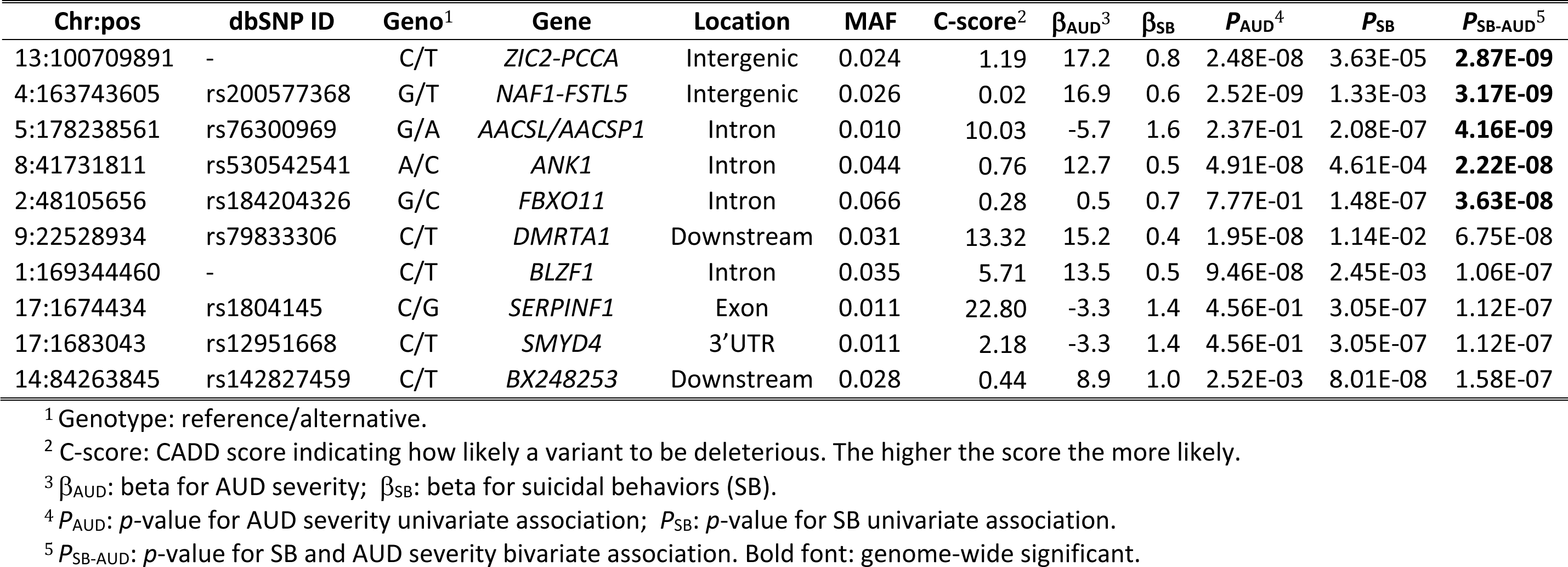
Top variants identified in bivariate genome-wide associations for suicidal behaviors and AUD severity in the American Indians.

| Chr:pos | dbSNP ID | Geno<sup>1</sup> | Gene | Location | MAF | C-score<sup>2</sup> | $\beta_{\text{AUD}}$<sup>3</sup> | $\beta_{\text{SB}}$ | $P_{\text{AUD}}$<sup>4</sup> | $P_{\text{SB}}$ | $P_{\text{SB-AUD}}$<sup>5</sup> |
| --- | --- | --- | --- | --- | --- | --- | --- | --- | --- | --- | --- |
| 13:100709891 | - | C/T | *ZIC2-PCCA* | Intergenic | 0.024 | 1.19 | 17.2 | 0.8 | 2.48E-08 | 3.63E-05 | **2.87E-09** |
| 4:163743605 | rs200577368 | G/T | *NAF1-FSTL5* | Intergenic | 0.026 | 0.02 | 16.9 | 0.6 | 2.52E-09 | 1.33E-03 | **3.17E-09** |
| 5:178238561 | rs76300969 | G/A | *AACSL/AACSP1* | Intron | 0.010 | 10.03 | -5.7 | 1.6 | 2.37E-01 | 2.08E-07 | **4.16E-09** |
| 8:41731811 | rs530542541 | A/C | *ANK1* | Intron | 0.044 | 0.76 | 12.7 | 0.5 | 4.91E-08 | 4.61E-04 | **2.22E-08** |
| 2:48105656 | rs184204326 | G/C | *FBXO11* | Intron | 0.066 | 0.28 | 0.5 | 0.7 | 7.77E-01 | 1.48E-07 | **3.63E-08** |
| 9:22528934 | rs79833306 | C/T | *DMRTA1* | Downstream | 0.031 | 13.32 | 15.2 | 0.4 | 1.95E-08 | 1.14E-02 | 6.75E-08 |
| 1:169344460 | - | C/T | *BLZF1* | Intron | 0.035 | 5.71 | 13.5 | 0.5 | 9.46E-08 | 2.45E-03 | 1.06E-07 |
| 17:1674434 | rs1804145 | C/G | *SERPINF1* | Exon | 0.011 | 22.80 | -3.3 | 1.4 | 4.56E-01 | 3.05E-07 | 1.12E-07 |
| 17:1683043 | rs12951668 | C/T | *SMYD4* | 3'UTR | 0.011 | 2.18 | -3.3 | 1.4 | 4.56E-01 | 3.05E-07 | 1.12E-07 |
| 14:84263845 | rs142827459 | C/T | *BX248253* | Downstream | 0.028 | 0.44 | 8.9 | 1.0 | 2.52E-03 | 8.01E-08 | 1.58E-07 |

<sup>1</sup> Genotype: reference/alternative.
<sup>2</sup> C-score: CADD score indicating how likely a variant to be deleterious. The higher the score the more likely.
<sup>3</sup> $\beta_{\text{AUD}}$ : beta for AUD severity; $\beta_{\text{SB}}$ : beta for suicidal behaviors (SB).
<sup>4</sup> $P_{\text{AUD}}$ : $p$ -value for AUD severity univariate association; $P_{\text{SB}}$ : $p$ -value for SB univariate association.
<sup>5</sup> $P_{\text{SB-AUD}}$ : $p$ -value for SB and AUD severity bivariate association. Bold font: genome-wide significant.

Variants associated with SB-AUD at *p* < 10^-6^ were mapped to 31 genes. The majority of these variants are intronic, upstream, or downstream from a gene. Rs1804145 on *SERPINF1* is the only nonsynonymous variant and has a C-score of 22.8 (top 0.52% most deleterious). These genes are enriched for 27 transcription factor (TF) targets and three microRNA targets (Table S3), suggesting that the top genes share certain regulatory motifs. These genes are most significantly up-regulated in artery, and down-regulated in areas included in the basal ganglia (nucleus accumbens, caudate, and putamen). In addition, they are significantly differentially expressed in anterior cingulate cortex, hypothalamus, substantia nigra, and kidney (Figure 2). Many psychiatric or neurological disorders have been linked to basal ganglia targets including addiction and depression (63).

**Figure 2.**
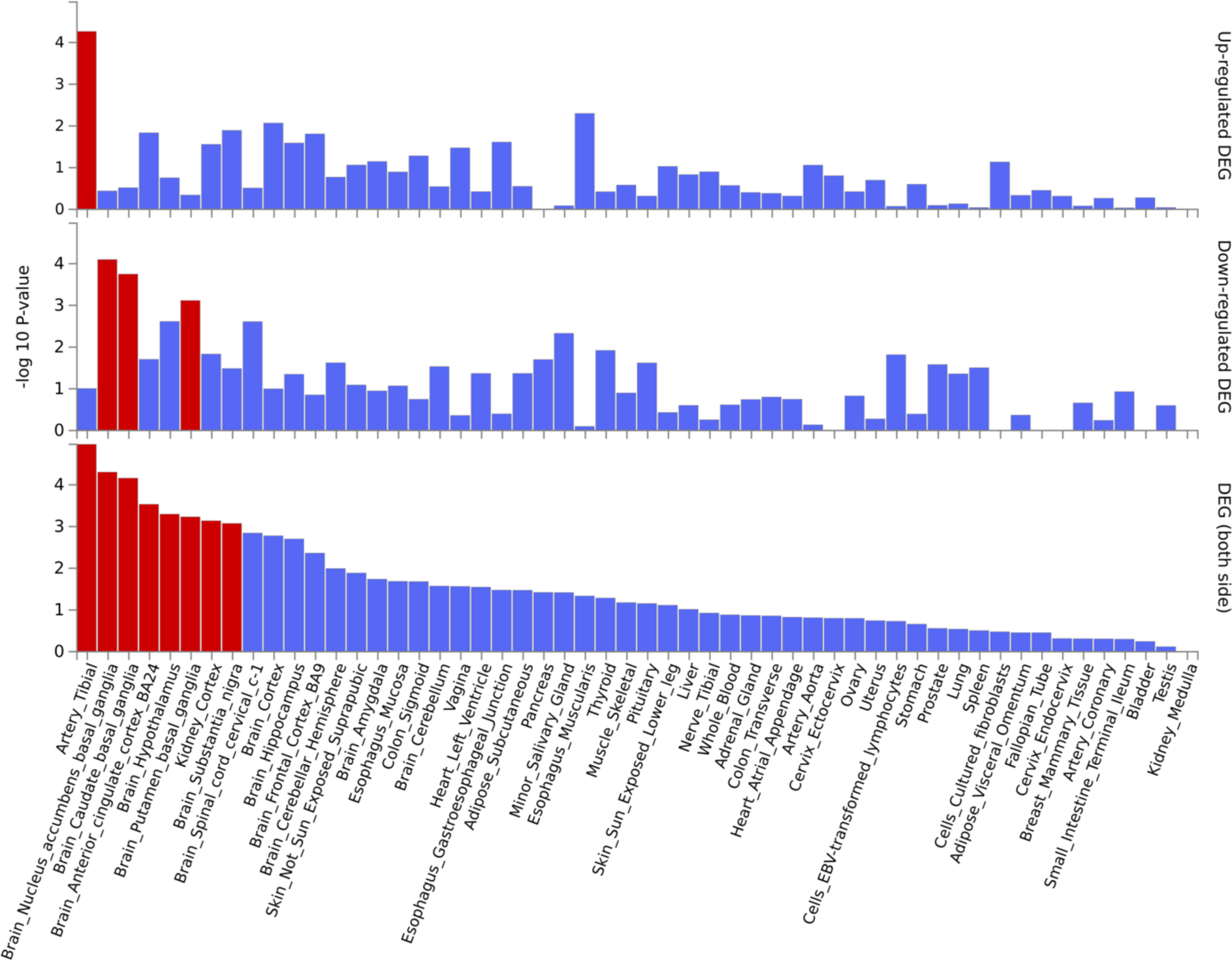
Enrichment in tissue-specific differentially expressed gene sets (DEG) of top genes associated with SB-AUD. The tissue-specific differential gene expression test was conducted against all genes across genomes that exhibited significantly increased or decreased expression levels in a certain tissue sample compared to all other samples. The analysis was performed using FUMA and utilized tissue-specific transcriptome data across 54 tissue types from GTEx v8. From top to bottom: up-regulated, down-regulated, differentially expressed. Red: Significantly enriched (*p* < 0.05 with Bonferroni correction).

### 3.2 Genes with rare and low-frequency variants associated with suicidal behaviors in AI

Rare variant analysis identified six genes and one long intergenic non-coding RNA (lincRNA) as being significantly associated with SB (FDR<0.05), and an additional four genes strongly associated with SB (FDR<0.1) (Table 2). *SERPINF1* was the top gene linked to SB. Both the *nonsyn* (*p*=1.3E-6) and the *exonreg* (*p*=1.7E-6) variant groups of this gene remained genome wide significant after a multiple testing correction. Four out of the nine *nonsyn* rare variants in *SERPINF1* had C-scores between 23 and 29, representing between 0.1-0.5% most deleterious mutations. One of the four probably damaging variants, rs1804145, was also suggestively associated with SB-AUD in the bivariate analysis (*p*=1.1E-7, see Table 1). Additional genes significantly associated with SB included: *ZNF30*, *CD34*, *SLC5A9*, *OPRD1*, and *HSD17B3*. A lincRNA *AC002511.3* is also associated with SB. Of the 15 *nonsyn* variants on *ZNF30*, one has a C-score of 33 (0.05% most deleterious), and six have C-scores between 22 and 24 (∼0.5% most deleterious). One of the nine *nonsyn* variants on *CD34* has a C-score of 32. Seven out of the 12 *nonsyn* variants on *SCL5A9* have C-scores ranging from 24 to 30. *OPRD1* has three rare exon variants, one of which has a C-score of 18 (1.6% most deleterious), and one of the 10 rare variants on *HSD17B3* has a C-score of 22.8 (∼0.5% most deleterious).

**Table 2.** Top genes with rare and low-frequency variants associated with suicidal behaviors in the American Indians.

| Chr | Positions | Genes<sup>1</sup> | Variants<sup>2</sup> | SNPs<sup>3</sup> | C-scores<sup>4</sup> | %Rare<sup>5</sup> | *p*-value<sup>6</sup> | FDR<sup>7</sup> |
| --- | --- | --- | --- | --- | --- | --- | --- | --- |
| 17 | 1673263-1680721 | **SERPINF1** | Nonsyn | 9 (10) | 0-4-0 | 5.5 | 1.32E-06 | **0.024** |
| 17 | 1665330-1680847 | **SERPINF1** | ExonReg | 12 (17) | 1-4-0 | 5.9 | 1.65E-06 | **0.024** |
| 19 | 35418193-35435632 | **ZNF30** | Nonsyn | 15 (21) | 3-6-1 | 8.5 | 1.76E-06 | **0.024** |
| 1 | 208062056-208073330 | **CD34** | Nonsyn | 9 (12) | 2-0-1 | 2.7 | 3.55E-06 | **0.032** |
| 19 | 35923858-35924757 | **AC002511.3** | ExonReg | 3 (5) | 0-0-0 | 2.6 | 3.89E-06 | **0.032** |
| 1 | 48694570-48713100 | **SLC5A9** | Nonsyn | 12 (16) | 1-7-0 | 8.7 | 6.44E-06 | **0.044** |
| 1 | 29138936-29190138 | **OPRD1** | ExonReg | 3 (7) | 1-0-0 | 3.1 | 7.44E-06 | **0.044** |
| 9 | 98997810-99064425 | **HSD17B3** | ExonReg | 10 (12) | 0-1-0 | 10.4 | 9.20E-06 | **0.047** |
| 1 | 48688376-48714188 | **SLC5A9** | ExonReg | 27 (35) | 3-7-0 | 20.2 | 1.03E-05 | **0.047** |
| 2 | 74588717-74598791 | *DCTN1* | Nonsyn | 11 (11) | 2-7-1 | 9.6 | 1.73E-05 | 0.072 |
| 15 | 81605656-81614811 | *STARD5* | Nonsyn | 3 (3) | 2-1-0 | 5.7 | 2.39E-05 | 0.087 |
| 10 | 33475282-33552763 | *NRP1* | Nonsyn | 9 (12) | 2-4-0 | 8.6 | 2.53E-05 | 0.087 |
| 19 | 46807185-46832554 | *HIF3A* | Nonsyn | 11 (14) | 1-5-0 | 14.1 | 2.89E-05 | 0.092 |

<sup>1</sup> Bold font: gene is significantly associated with suicidal behaviors at the genome level after multiple comparison correction.
<sup>2</sup> Variant groups: ExonReg includes variants on exon and upstream/downstream of a gene; Nonsyn includes nonsynonymous and splicing variants of a gene.
<sup>3</sup> Number of rare/low-frequency markers included in the test for each gene. The number in the parenthesis is the total number of SNPs of the same category on the gene.
<sup>4</sup> C-scores counts: three numbers c1-c2-c3 represent the number of rare variants that have CADD scores (C-scores) in the range of 15-20 (1-3% most deleterious mutations), 20-30 (0.1-1% most deleterious), and >30 (<0.1% most deleterious). For instance, the first row *SERPINF1* Nonsyn group has 9 variants. 0-4-0 indicates that 0 of the 9 SNPs has C-scores in the range of 15-20 or over 30, while 4 SNPs have C-scores between 20 and 30. The higher the C-score, the more likely a variant is deleterious.
<sup>5</sup> The fraction of individuals that have at least one of the rare/low-frequency markers on the gene.
<sup>6</sup> Nominal *p*-values.
<sup>7</sup> FDR-adjusted *p*-values, combining the two gene variant groups (Number of genes tested in each group: ExonReg = 28718; Nonsyn = 12588). Bold font: genome-wide significance (FDR<0.05).

A gene set enrichment analysis for the 11 genes whose rare and low frequency variants were associated with SB at FDR<0.1 (Table 2) identified *axon* to be the most significantly enriched gene ontology term (nominal *p*=1.63E-5, adjusted *p*=0.016). Four of the 11 genes belong to the axon gene set: *OPRD1*, *NRP1*, *SERPINF1*, and *DCTN1*. Network analysis conducted using GeneMania for the 11 genes in Table 2 recognized 10 genes and automatically selected 10 additional related genes (Figure 3). The analysis considered pathways and genetic interactions and identified a number of significantly associated functional networks (Table S4). Figure 3 illustrates the top enriched distinct functions, including VEGF signaling (FDR=2.1E-04), angiogenesis (FDR=2.1E-04), and response to decreased oxygen levels (FDR=4.2E-03). VEGF, as an agngiogenic cytokine, and hypoxia have been associated with depression and suicide (64, 65).

**Figure 3.**
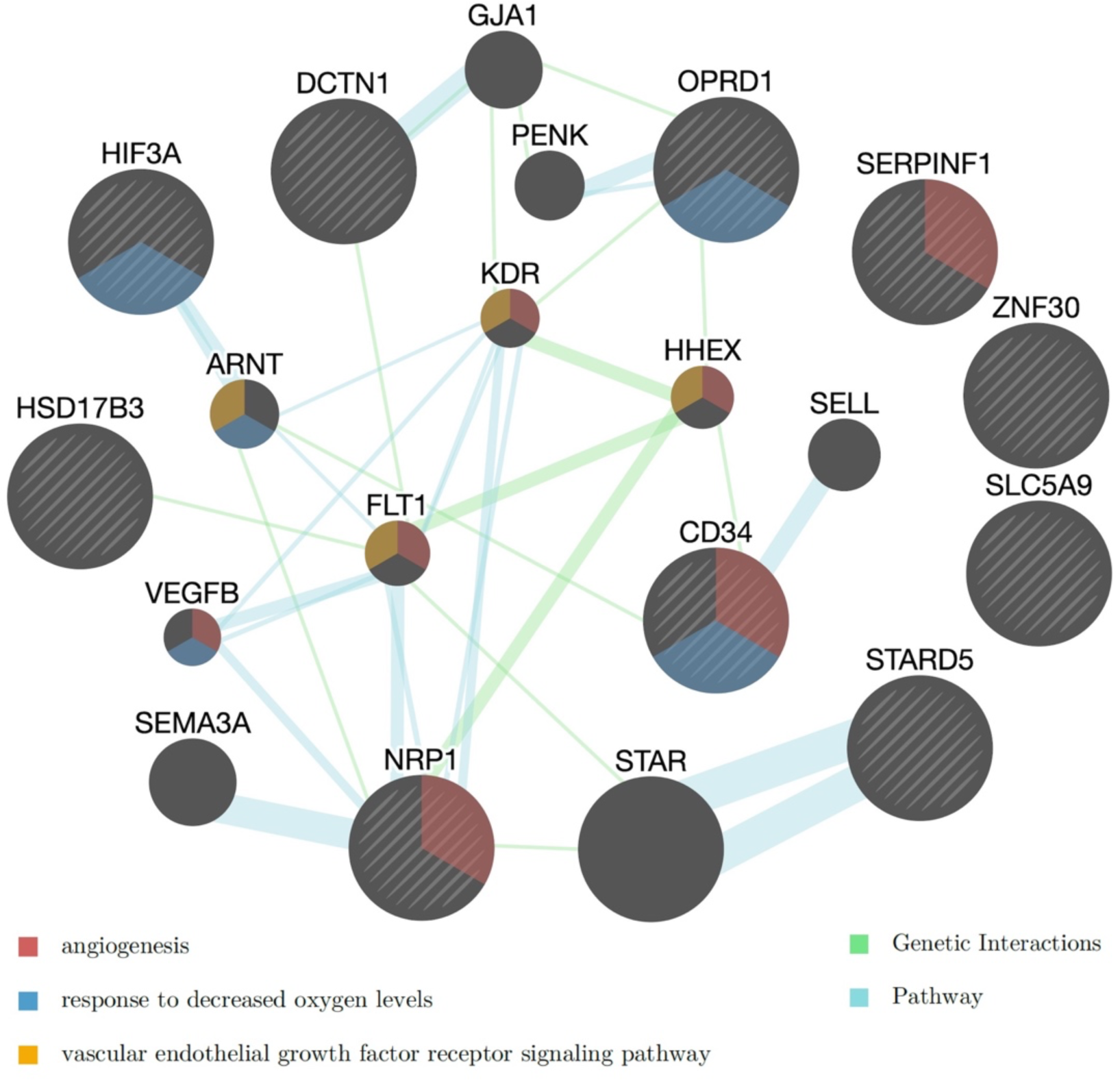
Networks of the rare-variant genes associated with suicidal behaviors in AI. Three of the enriched functional networks are highlighted in the gene nodes (see Table S4 for the complete list). Yellow: vascular endothelial growth factor (VEGF) receptor signaling pathway (FDR=2.1E-04); Red: angiogenesis (FDR=2.1E-04); Blue: response to decreased oxygen levels (FDR=4.2E-03). The color of the edges linking the genes indicate the type of database used in the GeneMania analysis. Blue: Pathways; Green: Genetic interactions.

### 3.3. Pathways with rare and low-frequency variants associated with suicidal behaviors in AI

Regulation of gene expression by hypoxia inducible factor (HIF) is the top pathway (*p*=1.4E-5) whose rare and low-frequency variants were associated with SB (Table 3). The pathway is comprised of 10 genes with total of 91 nonsynonymous or splicing site variants (*nonsyn*), of which 83 have MAF<5% and were included in the association test. Hypoxia could decrease serotonin synthesis, which has been linked to suicide (66). The three additional pathways that were associated with SB at FDR<0.1 are essentially two distinct pathways: vasopressin regulated water reabsorption with 164 *nonsyn* rare variants on 44 genes, and cellular hexose transport with 126 *nonsyn* rare variants on 21 genes. Vasopressin plays a critical role in regulating renal water reabsorption and cardiovascular homeostasis. As an integral part of the hypothalamic-pituitary-adrenal (HPA) axis, it is also an important player in response to stress and contributes to stress-related disorders such as anxiety and depression (67). The cellular hexose transport pathway includes gene families responsible for glucose transport in humans. Sodium independent glucose transporters (GLUTs) are encoded by the SLC2 family, while sodium dependent glucose transporters (SGLTs) are encoded by the SLC5 family.

**Table 3.**
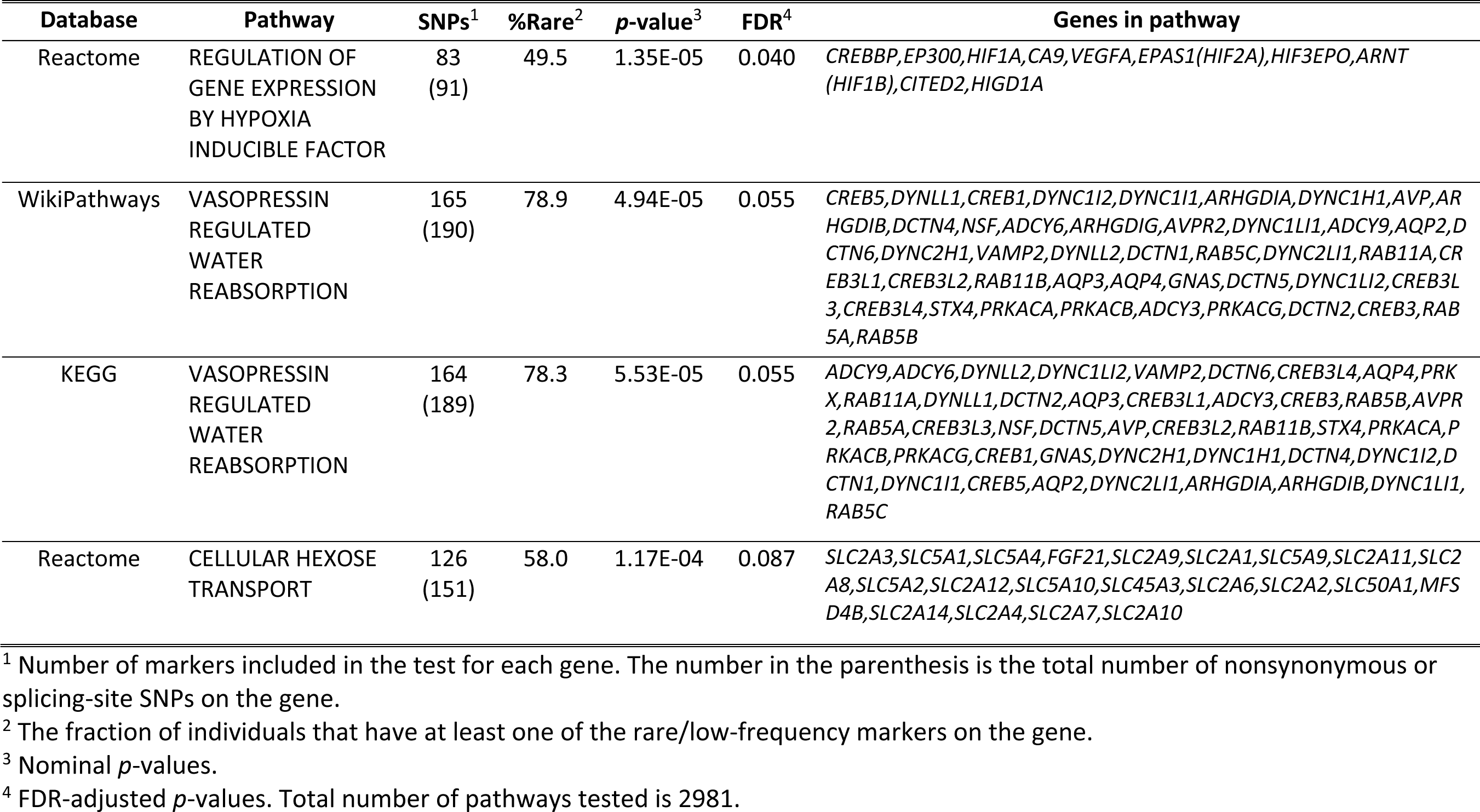
Top pathways or gene sets with rare and low-frequency nonsynonymous variants associated with suicidal behaviors (FDR<0.1) in the American Indians.

## 4. DISSCUSSION

Despite unprecedented strategies to support its prevention as a public health imperative, suicide has, alarmingly, increased in recent years—now representing the second leading cause of death in teens and young adults in the U.S. as well as worldwide (68, 69). American Indians and Alaska Natives (AI/AN) demonstrate the highest burden of suicide compared to all other ethnic or racial groups, highlighting urgency to critically advance research and prevention (3–6).

However, rates of suicide vary substantially between tribal groups and by distinct geographic region (2), underscoring the need to delineate risk and resilience factors in local communities to develop community-specific prevention and intervention efforts (10–12, 70). The present study focused on an American Indian population living on eight contiguous reservations. This population demonstrates high rates of suicide as well as AUD (19, 43). In this study, we investigated potential genetic risk factors for suicidal behaviors by studying: (1) the possible genetic overlap of SB with AUD, and (2) the impacts of rare- and low-frequency genomic variants on SB. We identified five variants significantly associated with SB-AUD, two of which are intergenic and three are intronic. Two of the five variants, although not significantly associated with either SB or AUD severity on their own in this AI cohort, were significantly associated with the dual phenotype SB-AUD—suggesting the genetic correlations were induced by residual shared factors. Six genes, one lincRNA, and one canonical pathway, were found to be significantly associated with SB through rare and low-frequency variant analysis. Four additional genes, and two pathways, also were strongly associated with SB.

### F-box gene *FBXO11* significantly associated with SB-AUD in this AI population

The intronic variants significantly associated with SB-AUD are on genes *AACSP1*, *ANK1*, and *FBXO11*. *AACSP1* is a pseudogene whose function is unclear, although it has been associated with an Alzheimer’s marker in an interaction analysis (71). *ANK1* encodes ankyrin-1 protein active in red blood cells as well as brain and muscle. The gene has been linked to Alzheimer’s disease through epigenetic deregulation (72, 73) and appears to play a role in immunomodulation (74).

Rs184204326 on gene *FBXO11* was the only common variant significantly associated with SB-AUD. *FBXO11* encodes a member of the F-box protein family and is highly conserved in evolution. It is part of the SCF (SKP1-cullin-F-box) complex, which is responsible for ubiquitination and degradation of SCF substrates and plays an important role in the maintenance of genome stability (75). *FBXO11* is also involved in regulating alternative splicing (76). While the gene is expressed in various tissues, it is particularly abundant in the brain. *FBXO11* has not been previously associated with SB. However, variants on or near *FBXO11* have been associated with a number or correlated behavior traits and disorders such as alcohol consumption (77), smoking initiation (77, 78), risk taking behaviors (79, 80), externalizing behaviors (81), educational attainment (82), insomnia (83), schizophrenia and depression (84, 85). *De novo* mutations on *FBXO11* have also been found to cause intellectual disability with behavior problems as well as facial dysmorphisms (86) and other neurodevelopmental disorders (87). Variant rs77969729 on *FBXO11* has been linked to Alzheimer’s disease in the UKBiobank (88). This variant is in LD with rs184204326 on *FBXO11* in the AI cohort and associated with SB-AUD at *p*=3.4E-7.

### Rare mutations in PEDF are likely a risk factor for suicidal behaviors

Six genes and one lincRNA were found to be significantly associated with SB through rare variant analysis. The top gene *SERPINF1* encodes serpin F1, also known as pigment epithelium-derived factor (PEDF). This gene has nine rare nonsynonymous or splicing site mutations, of which four are among the 0.1-0.5% most deleterious. Nearly 6% of individuals in the AI cohort carry some of these mutations. PEDF is a secreted glycoprotein and a potent inhibitor of angiogenesis (89). It also has neuroprotective effects and been implicated in depression (90, 91). Reduction in PEDF levels have been found in the plasmas of patients with major depressive disorder (MDD), as well as in the prefrontal cortex (PFC) of animal models exhibiting depressive-like behaviors (91). Conversely, overexpression of PEDF in the PFC has been shown to induce antidepressant “like” behaviors, by exerting effects on the tryptophan and glutamate in the PFC, with tryptophan being an essential amino acid precursor of serotonin, which is known to be associated with both depression and SB (92). Another study has shown that PEDF in the hippocampus has a similar effect on depressive phenotypes in animal models by contributing to the synaptic formation and Wnt signaling activation in that region (93). PEDF has thus been suggested as a biomarker and a novel therapeutic target for depression (90).

Additional genes significantly associated with SB in the AI cohort included *ZNF30*, *CD34*, *SLC5A9*, *OPRD1*, and *HSD17B3*. *ZNF30* encodes a zinc finger protein. A microdeletion of five genes including *ZNF30* results in chromosome 19q13.11 deletion syndrome that includes features such as: developmental delay, microcephaly and intellectual disabilities (94). The GWAS meta-analysis by the international suicide genetics consortium (ISGC) reported another zinc finger family gene (*ZNF28*) associated with SB and suicide death (31). *CD34* is involved in the innate immune system. Its variants have been associated with Alzheimer’s disease (88) and risk-taking behavior (80). *SCL5A9* is involved in sodium ion transport and is also known to be a sodium-dependent glucose transporter (95). This gene has been associated with amygdala volume (96) and metabolic measurements (97). Variants on or near *HSD17B3* have been linked to smoking behavior (98), brain morphology measurements (99), memory performance (100) and Alzheimer’s markers (71, 101).

Opioid receptor delta 1 (*OPRD1*) gene encodes a member of the opioid family of the G-protein coupled receptors (GPCR). Delta opioid receptors are involved in reward mediation and neuroprotection. *OPRD1* is specifically involved in the opioid receptor signaling pathway and cellular response to hypoxia. Numerous candidate gene studies have implicated *OPRD1* in addiction, including opioid, cocaine, and alcohol dependence (102, 103). Variants on *OPRD1* have also been associated with schizophrenia (104) and educational attainment (82).

### Hypoxia regulation is significantly associated with suicidal behaviors

*Nonsyn* rare variants in a pathway related to hypoxia inducible factor (HIF) regulation were significantly associated with SB (see Table 3). Nearly half of the individuals in the AI cohort have some of these rare mutations on their genes in this pathway. The network analysis of the top rare-variant genes associated with SB also found that those genes were enriched for response to decreased oxygen levels (see Figure 3).

Chronic hypoxia is suggested to be a risk factor for suicide (64), and metabolic stress associated with hypoxia is a possible mechanism (105). Hypoxia is also hypothesized to increase the risk of suicide by reducing the synthesis of brain serotonin (66) or downregulating PEDF (106), which has a protective role in depression and is associated with SB in this AI cohort. HIFs are transcriptional factors that respond to reduced oxygen levels in cell and tissue. HIF-1 protects against hypoxia and reduces oxidative stress (107), while HIF-2α plays an important in the modulation of inflammatory responses (108). Recent studies have indicated that oxidative stress and abnormal energy metabolism in the brain play significant roles in the development of depression. Therefore, increasing HIF-1 activity has been suggested as a potential new therapeutic target for depression and suicide ideation (107). Gene expression analyses have demonstrated that patients with MDD exhibit increased expression of HIF-1 and its target genes, including *VEGF* and *GLUT1* (*SLC2A1*) (65). Our network analysis of the top rare-variant genes associated with SB in the AI have also found enrichments in VEGF receptors signaling and angiogenesis.

Alcohol exposure also can alter expression of HIF. For instance, alcohol exposure can induce HIF-1α activation, however the dose and timing of alcohol exposure results in differential expression of HIF-1α in the brain and other organs (109). Both acute and chronic alcohol exposure have been found to increase HIF-1α expression in the brain cortex, whereas chronic binge alcohol exposure decreased HIF-1α expression (109). Another study found HIF3A can be epigenetically induced in the amygdala in animal models by acute alcohol exposure, and its epigenetic reprogramming was associated with the anti-anxiety effect of acute alcohol exposure (110). *Nonsyn* rare variants in *HIF3A* were strongly associated with SB (nominal *p*=2.9E-05, FDR=0.09) in the present study.

### Vasopressin regulated water metabolism and hexose transport strongly associated with suicidal behaviors

*Nonsyn* rare variants in two additional pathways, vasopressin-regulated water reabsorption and cellular hexose transport, were found to be strongly associated with SB in the AI (FDR<0.1). Vasopressin is an evolutionary ancient neuropeptide that is involved in regulating physiological processes such as renal water reabsorption and cardiovascular homeostasis. It also plays an important role in the modulation of emotional and social behaviors in the brain (111, 112), with vasopressin containing neurons most abundantly found in the hypothalamus. The vasopressin system is known to interact with the HPA axis (67). Indeed, cortisol response and stress reactivity within the HPA axis is well-established as an endophenotype for depression and SB, as increased cortisol level is associated with death by suicide (113, 114). HPA axis dysfunction has also been observed in those with a history of suicide attempts (114). In addition, changes in water and electrolyte metabolism have been reported in clinical studies of depressed patients (115, 116). Alcohol also uniquely interacts with the vasopressin system. Alcohol is a diuretic that promotes water loss by inhibiting the production of vasopressin. The HPA axis response to alcohol can be altered by manipulating the vasopressin system (117). The vasopressin system has thus become an emerging therapeutic target for stress and depression, as well as alcohol-related behaviors (67, 117). Gene families in the cellular hexose transport pathway mediate glucose absorption in the small intestine, glucose reabsorption in the kidney, glucose uptake by the brain across the blood-brain barrier, and glucose release by all cells in the body (118). Evidence suggests that disturbances in glucose metabolism may be associated with suicidal ideation and attempts (119). A recent study has found a significant association between blood glucose and suicide attempts in male patients with MDD (120). Taken together, our findings suggest that pathways underlying vasopressin-regulated water reabsorption and the cellular hexose transport system warrant additional investigation in association with SB.

### Strengths and Limitations

The results of this study should be interpreted in light of several limitations. The analyses were not meant to generate a comprehensive model of suicide risk and AUD in this community group, but rather to determine whether specific genetic associations could be identified for suicidal thoughts and acts, and between SB and AUD phenotypes using genetic analyses. A larger sample, powered to access and assess additional variables associated with suicide risk, particularly according to the above systems, is recommended. Next, our findings may not generalize to other American Indians in the population from which the sample was drawn or be representative of all American Indians, as rates of AUD and suicide vary among tribes (17, 19, 121). In addition, we used retrospective data for lifetime measures of suicide risk, which are subject to recall bias and may include reporting bias for psychiatric symptoms. Lifetime suicidal ideation and behaviors were assessed using a subscale of the SSAGA as well as verified death records. Prospective investigations, using validated measures of suicidal symptom severity and intensity of suicidal ideation, and indexing the severity and lethality of SB, are recommended to further elucidate specific symptom relationships. Despite these limitations and in consideration of the current sample size, we were able to identify several genes and pathways implicated in SB and AUD severity. Our findings suggest that rare variant analysis in a high-risk population, enabled by whole-genome sequencing, has the potential to identify novel genetic factors influencing suicidal behaviors. Bivariate association analysis between comorbid disorders may further increase statistical power when the same loci potentially contribute to both disorders.

In conclusion, we conducted the first genome-wide bivariate association analysis for SB and AUD, and identified five genome-wide significant loci. We also conducted the first large-scale rare variant analysis and identified 11 novel genes and three pathways for SB. Of particular importance, this study represents the first investigation of genetic factors for SB in an American Indian population that has high risk for suicide. Although our findings may be population specific, the rare functional mutations relating to PEDF and HIF regulation may suggest an important mechanism underlying suicidal behaviors and a potential therapeutic target for treatment and intervention in the prevention of suicide.

## Data Availability

The data that support the finding of this study are available by contacting the last author. However, data availability is subject to approval of the specific AI tribes participating.

## Ethical standards

The authors assert that all procedures and protocols for the study were carried out in accordance with the latest version of the Declaration of Helsinki. The protocol and procedures were approved by the Institutional Review Board of The Scripps Research Institute and Indian Health Council for the AI tribes participating. Written informed consent was obtained from each participant after the study was fully explained by study staff.

## Role of the funding source

This work was supported by the National Institutes of Health (NIH): National Institute on Alcohol Abuse and Alcoholism (NIAAA) R01AA026248 and R01AA027316 to CLE and K25AA025095 to QP, and National Institute on Drug Abuse (NIDA) DP1DA054373 to QP. NIAAA and NIDA had no further role in study design; collection, analysis, and interpretation of data; writing of the report; or the decision to submit the article for publication. The authors report no conflicts of interest, competing interests or undisclosed financial support. The authors alone are responsible for the content and writing of this paper, and the views expressed do not necessarily represent the views of the NIH, NIAAA or NIDA.

## Author Contributions

**Qian Peng:** Conceptualization, Methodology, Software, Formal analyses, Validation, Writing - original draft, review & editing, Visualization, Funding acquisition. **Cindy L Ehlers:** Data acquisition, Conceptualization, Methodology, Formal analyses, Writing - original draft, review & editing, Supervision, Project administration, Visualization. **Rebecca Bernert:** Conceptualization, Methodology, Writing-review & editing, **David A. Gilder:** Supervision, Formal analysis, Data curation, Writing-review & editing, **Kate Karriker-Jaffe**: Conceptualization, Project administration, Supervision, Writing-review & editing.

## Notes

### Competing Interest Statement

The authors have declared no competing interest.

### Author Declarations

IRB of The Scripps Research Institute gave ethical approval for this work.

